# Topological changes of fast large-scale brain dynamics in Mild Cognitive Impairment predict the decay of the hippocampal memory

**DOI:** 10.1101/2022.11.11.22282206

**Authors:** Antonella Romano, Emahnuel Troisi Lopez, Lorenzo Cipriano, Marianna Liparoti, Roberta Minino, Arianna Polverino, Carlo Cavaliere, Marco Aiello, Carmine Granata, Giuseppe Sorrentino, Pierpaolo Sorrentino

## Abstract

Functional connectivity has been widely used as a framework to investigate widespread brain interactions underlying cognitive deficits in Mild Cognitive Impairment (MCI). However, one of the main constraints of functional connectivity is that it is averaged over a time interval and therefore may not take into account the aperiodic and scale-free burst of activity (i.e., the neuronal avalanches) characterising the large-scale dynamic activity of the brain. Here, we used the recently proposed Avalanche Transition Matrix framework to source-reconstructed magnetoencephalography signals in a cohort of 32 MCI patients and 32 healthy controls (HC) to deepen the spatio-temporal features of neuronal avalanches and explore their topological properties. Our results showed that MCI patients exhibited a more centralised network (as assessed by higher values of degree divergence and leaf fraction) compared to HC. Furthermore, we found that the degree divergence (in the theta band) was predictive of the episodic memory impairment, assessed by FCSRT immediate total recall. These findings highlight the role of dynamical features in detecting functional and structural changes in clinical conditions. Hopefully, the proposed framework may be helpful in monitoring the development of the disease by adding subtle information that contributes to a more thorough phenotypical assessment of patients.

## 1. INTRODUCTION

Mild cognitive impairment (MCI) is a preclinical, transitional stage between healthy ageing and dementia. Its prevalence ranges from 6.7% to 25.2% in people older than 60 years old and increases with age and lower education level. Nowadays, MCI is considered as a timeframe during which we should act to delay conversion to dementia (Jongsiriyanyong and Limpawattana, 2018). From a clinical standpoint, MCI patients are classified according to type and number of affected cognitive domains. The first distinction is made on the presence/absence of memory impairment, giving rise to the two major subtypes of MCI, namely amnestic (aMCI) and non-amnestic (naMCI), where the former denotes a memory loss, while the latter refers to a cognitive impairment which, by sparing memory, mainly affects specific cognitive domains such as executive functions, attention, visuospatial ability or language. Importantly, aMCI is considered as the subtype most likely to protend a diagnosis of Alzheimer’s disease (AD) (Buldú et al., 2011; Celone et al., 2006). Noteworthy these clinical forms may evolve over time, so if more than one domain is involved (in both aMCI and naMCI) the label of “multiple-domain”, in opposition to the “single-domain”, is used. (Petersen, 2016). For example, as the aMCI progresses, other cognitive functions may become compromised, defining a continuum between the amnestic and the multiple domain aMCI. Hence, it is highly misleading to constrain the cognitive deficits in MCI to the malfunctioning of any specific brain region, rather they stem from the miss-interactions among multiple brain regions (Liu et al., 2012; Minati et al., 2014; Sorrentino et al., 2018; Sorrentino, et al., 2021).

Network theory has been used as a framework to characterise large-scale interactions, in order to pinpoint specific mechanisms underpinning cognitive deficits. To this end, Functional Connectivity (FC), which typically measures the pair-wise statistical dependencies between regional signals (Friston, 2011), has been applied in health and disease (Fornito et al., 2015). The FC changes reported in MCI were non-homogeneous even within individual studies, as the strength of the connectivity was increased in some brain regions and decreased in others (Contreras et al., 2017; López-Sanz et al., 2017; Jacini et al., 2018; Liu et al., 2012; López et al., 2017). Moreover, MCI subjects display a hypersynchronised network, which then desynchronises if overt disease sets in (Buldú et al., 2011; Celone et al., 2006). Importantly, the reported FC changes failed to convincingly replicate across studies, and new approaches are needed to provide reliable estimates of large-scale brain interactions.

A source of such variability could arise from a bias in most functional connectivity (FC) studies which generally assume a stationary condition in brain activity. These studies, by focusing on “average” connectivity over a time interval, disregards the evolution of the coactivations over time (i.e., brain dynamics) (Zalesky et al., 2014). In fact, it was shown that large-scale brain activity is far from stationary, and instead it is characterised by aperiodic, scale-free bursts of activity (in the context of statistical mechanics often referred to as neuronal avalanches), which are indeed expected, given a nonlinear underlying dynamics (Haldeman & Beggs, 2005; Tagliazucchi et al., 2012, Shriki et al., 2013). Hence, the study of time averaged connectivity might not be optimal to capture this bursty, non-linear activity.

The dynamics of a healthy brain has been shown to display high flexibility, since the regions that are recruited change at each subsequent avalanche, generating complex spatio-temporal patterns. Hence, the number of avalanche patterns, i.e., the functional repertoire, has been used as a proxy for the flexibility of brain activity. Flexible dynamics is maintained in health and, conversely, is disrupted in disease (Muñoz, 2018; Sorrentino et al., 2021). In fact, smaller functional repertoires occur in neurological diseases and are predictive of both clinical disability and disease progression (Polverino et al., 2022; Sorrentino et al., 2021).

In this study, in analogy with previous findings (Polverino et al., 2022; Sorrentino et al., 2021), we hypothesise that MCI would result in a more stereotyped brain dynamics, as measured by a smaller functional repertoire. Then we moved on to an in-depth characterization of the spatio-temporal dynamics of avalanches (Sorrentino et al., 2021). In fact, perturbations of local activation do not occur randomly, rather, they spread preferentially across the white-matter bundles (i.e., the structural connectome) (Sorrentino et al., 2021). As a consequence, not all regions are recruited equally by the avalanches (as the topology of the structural connectomes will tend to attract the functional dynamics to its eigenmodes) (Tewarie et al., 2022). We hypothesise that the reduction in the flexibility of brain dynamics would be mirrored by a rearrangement of the spreading of the perturbations on the large-scale activity. In particular, we predict that in MCI the regions that are structurally more central would be recruited more often as compared to healthy controls (which achieve a more flexible dynamics). Conversely, more peripheral regions would be recruited even less often in MCI subjects. The reasoning behind this hypothesis lies in the fact that in the healthy brain the spreading of neuronal avalanches is influenced by the structural connectome, but it is not fully determined by it. In fact, healthy subjects display a flexible dynamic that varies over time and ‘detaches itself’ from the structural connectome. In MCI, the lack of flexibility would reduce the number of dynamical reconfigurations, which would result in an overall stronger influence of the underlying structure over the spatio-temporal evolution of neuronal avalanches. Globally, this would result in changes in a rearrangement of the functional topology toward a more “centralised” network.

To test our hypothesis, we performed magnetoencephalography (MEG) recording on thirty-two MCI patients and thirty-two healthy controls. Then, the source-reconstructed brain signals were analysed to extract neuronal avalanches, operationally defined as an event starting when large fluctuations of activity are present in at least one brain region and ending when all the regions return to their baseline. Hence, we could consider an avalanche pattern as the set of brain regions recruited in each individual avalanche. In turn, the totality of the unique avalanche pattern (i.e., discarding repetitions) occurring over time delineates the functional repertoire, and the number of such patterns defines its size, which is used to quantify flexibility. Finally, the topological features of brain dynamics were assessed using the recently developed Avalanche Transition Matrix (ATM). The ATMs convey the spatio-temporal trajectories of neuronal avalanches as they spread across the brain and define the probability that two brain regions will move away from their baseline activity (i.e., avalanches are occurring) in two consecutive time epochs (Sorrentino et al., 2021). Finally, in order to verify a possible relationship between the topological organisation of brain dynamics in MCI and its clinical features, we built a multilinear regression model to test if the topological properties of the ATMs can predict the clinical impairment.

## 2 MATERIALS AND METHODS

### 2.1 Participants

Thirty-two MCI patients (18 males and 14 females; 21 single domain aMCI and 11 multiple domain aMCI; mean age 71.31; SD ± 6.83; mean education 10.54; SD ± 4.33) were recruited from the Centre of Cognitive and Memory Disorders of the Hermitage Capodimonte Clinic in Naples, Italy. All the patients were right-handed and native Italian speakers. The MCI diagnosis was done according to the National Institute on Ageing-Alzheimer Association (NIA-AA) criteria (Albert et al., 2011). Inclusion criteria were: (1) the absence of neurological or systemic illness that could affect the cognitive status, (2) no contraindications to MRI or MEG recording and (3) Fazekas score (for both periventricular white matter and deep white matter scores) ≤ 2. Thirty-two subjects (19 males and 13 females) matched for age (69.9±5.61) and education (12.96±4.56) were enrolled as a control group (HC). The cohort characteristics are summarised in table 1. Both HC and MCI patients underwent neurological examination, MRI scan (which included the estimation of the hippocampal volumes) and MEG recording. The neuropsychological screening (see table 2) included also the Free and Cued Selective Reminding Test (FCSRT) which is highly sensitive in detecting hippocampal mnestic deficits (Auriacombe et al., 2010; Grober & Buschke, 1987). In particular, the FCSRT assesses the ability of encoding and retrieval through semantic cues by exploiting and maximising the learning effect (Dubois et al., 2014). The first part of the test is defined as “immediate recall” and it aims to induce the semantic encoding while the second part (performed after 20 minutes) is defined as “delayed recall”. For both parts of the tests, the recall was first free and then cued (Auriacombe et al., 2010).

**Table 1:** Subjects characteristics. HC: healthy controls; MCI: Mild Cognitive Impairment; SD: Standard Deviation; NS: not significant. The structural information was available for 22 out of 32 healthy controls and for 30 out 32 MCI patients.

| **Parameters** | ***HC*** | ***MCI*** | ***Group differences*** |
| --- | --- | --- | --- |
| **Sample size (total)** | ***n 32*** | ***n 32*** | ***p value*** |
| **Age (mean ± SD)** | 69.9 (±5.61) | 71.31 (± 6.83) | NS |
| **Male/Female (n; %)** | 19/13<br>(59,37%; 40,62%) | 18/14<br>(56,25%;43,75%) | / |
| **Education level (mean ± SD)** | 12.96 (±4.56) | 10.54 (±4.33) | NS |
| **Sample size (MRI)** | ***n 22*** | ***n 30*** |  |
| **Left Hippocampal volume cm<sup>3</sup> (± SD)** | 3.37 (±0.35) | 2.81 (±0.55) | <0.001 |
| **Right Hippocampal volume cm<sup>3</sup> (± SD)** | 3.38 (± 0.33) | 2.82 (±0.43) | <0.001 |
| **Total Hippocampal volume cm<sup>3</sup> (± SD)** | 6.76 (±0.67) | 5.64 (±0.95) | <0.001 |

**Table 2:** Neuropsychological evaluation: MMSE: Mini Mental Score Examination (Folstein et al., 1975); FAB: Frontal Assessment Battery (Ilardi et al., 2022); BDI: Beck Depression Inventory(Sica & Ghisi, 2007); MDB: Mental Deterioration Battery (Carlesimo et al., 1996); FCSRT: Free and Cued Selective Reminding Test (Frasson et al., 2011); SD: Standard Deviation; NS: Not significant

| *Test* | *HC (n 32)*<br>*Mean (±SD)* | *MCI (n 32)*<br>*Mean (±SD)* | *p value* |
| --- | --- | --- | --- |
| **MMSE** | 27.56 (±1.69) | 26.4 (±1.72) | 0.012 |
| **FAB** | 16.21(±1.27) | 15.93 (±2.06) | NS |
| **BDI** | 8.06 (±4.41) | 9.43 (± 6.45) | NS |
| **MDB** |  |  |  |
| *Rey's 15-word immediate recall* | 44.85 (±6.05) | 26.54 (±5.89) | < 0.01 |
| *Rey's 15-word delayed recall* | 10.32 (±2.49) | 4.11 (±1.62) | < 0.01 |
| *Word fluency* | 39.32(±10.39) | 29.16 (±9.43) | < 0.01 |
| *Phrase construction* | 18.24 (±5.93) | 16.78 (±6.34) | NS |
| *Raven's 47 progressive matrices* | 28.22 (±3.98) | 24.25 (±5.53) | < 0.01 |
| *Immediate visual memory* | 19.71 (±1.71) | 17.91 (±3.25) | < 0.01 |
| *Freehand copying of drawings* | 10.02 (±1.24) | 9.19 (±2.07) | NS |
| *Constructive apraxia with landmarks* | 67.75 (±3.82) | 66.25 (±3.39) | NS |
| **FCSRT** |  |  |  |
| *FCSRT immediate free recall* | 30.36 (±2.83) | 22.34 (± 7.55) | < 0.001 |
| *FCSRT immediate total recall* | 35.92 (±0.26) | 32.21 (± 4.68) | < 0.001 |
| *FCSRT delayed free recall* | 10.15 (±1.11) | 5.13 (±3.58) | < 0.001 |
| *FCSRT delayed total recall* | 11.93 (±0.25) | 9.06 (±3.46) | < 0.001 |
| *FCSRT index of sensitivity of cueing* | 0.99 (±0.03) | 0.77 (± 0.22) | < 0.001 |

The study protocol was approved by the ‘‘Comitato Etico Campania Centro’’ (Prot.n.93C.E./Reg. n.14-17OSS) and all participants provided written informed consent in accordance with the Declaration of Helsinki.

### 2.2 MRI acquisition

MR images of both MCI patients and HC were acquired using a 3T Biograph mMR tomograph (Siemens Healthcare, Erlangen, Germany) equipped with a 12 channels head coil. The MR registration protocol was: (i) three-dimensional T1- weighted Magnetization-Prepared Rapid Acquisition Gradient-Echo sequence (MPRAGE, 240 sagittal planes, 214 × 21 mm2 Field of View, voxel size 1 × 1 × 1 mm3, TR/TE/TI 2400/2.5/1000 ms, flip angle 8°); (ii) Three-dimensional T2- weighted Sampling Perfection with Application optimised. Contrasts using different flip angle Evolution sequence (SPACE, 240 sagittal planes, 214 × 214 mm2 Field of View, voxel size 1 × 1 × 1 mm3, TR/TE 3370/563); (iii) Two-dimensional T2- weighted turbo spin echo Fluid Attenuated Inversion Recovery sequence (FLAIR, 44 axial planes, 230 × 230 mm2 Field of View, voxel size 0.9 × 0.9 × 0.9 mm3, TR/TE/TI 9000/95/25,00, flip angle 150°). The FreeSurfer software (version 6.0) (FreeSurfer, 2012) was used to obtain the volumetric analysis. Specifically, the hippocampal volumes were normalised by the estimated total intracranial volume (eTIV), while the Fazekas scale was used to evaluate the vascular burden (Fazekas et al., 1987). Two patients and ten healthy controls refused or did not complete the MR scan, and a standard MRI template was used instead.

### 2.3 MEG acquisition and pre-processing

Data were acquired using a MEG system composed of 154 magnetometers SQUID (superconductive quantum interference device) and 9 reference sensors. The acquisition took place in a magnetically shielded room (ATB, Biomag, ULM, Germany) to reduce external noise. To define the position of the head under the helmet we used Fastrack (Polhemus®), that digitised the position of four anatomical landmarks (nasion, right and left pre-auricular points and vertex of the head) and the position of four reference coils (attached to the head of the subject). Each subject was recorded twice (3.5 min each) with a one-minute break, in resting state with closed eyes. We also recorded the cardiac activity and the eyes movement to remove physiological artefacts. After applying an anti-aliasing filter, data were sampled at 1024 Hz. Data preprocessing was performed similarly to Liparoti et al. (Liparoti et al., 2021). Briefly, the MEG data were filtered in the band 0.5-48 Hz through the implementation of a 4^th^-order Butterworth IIR band-pass filter, using the Fieldtrip toolbox in the MATLAB environment. Moreover, a Principal Component Analysis (PCA) was carried out to reduce environmental noise. Finally, we performed a supervised Independent Component Analysis (ICA) to remove physiological artefacts from the ECG (one component per participant) and the EOG (no component per participant, rarely one) (Romano et al., 2022; Rucco et al., 2022)

### 2.4 Source reconstruction

The time series of the region of interest (ROIs), based on the Automatic Anatomical Labelling (AAL) Atlas (Gong et al., 2009), were reconstructed using the volume conduction as in (Nolte, 2003) and the Linearly Constrained Minimum Variance (LMCV) (Van Veen et al., 1997) beamformer algorithm, based on the native MRI of each subject. The time series were filtered in the five canonical frequency bands: delta (0.5 – 4 Hz), theta (4 – 8 Hz), alpha (8 – 13 Hz), beta (13 – 30 Hz), and gamma (30 – 48 Hz).

### 2.5 Analysis of brain dynamics

#### 2.5.1 Neuronal Avalanches and branching parameter

We estimated the “Neuronal Avalanches’’ to quantify the spatio-temporal fluctuations of brain activity. A neuronal avalanche is defined as an event starting when massive fluctuations of brain activity are present in at least one ROI and ending when all the ROIs return to their usual activity (Sorrentino et al., 2021). Firstly, we calculated the z-score of each ROIs’ time series, then each time series was thresholded according to a cut-off of 3 standard deviations (i.e., z = |3|). Note that we also changed the threshold from 2.5 to 3.5 to confirm that the results were not dependent upon the choice of the threshold (see supplementary materials 1). To be sure that we were actually capturing the critical dynamics (if present), we binned the time series. Similarly to Sorrentino et al., (Sorrentino et al., 2021) we estimated the suitable time bin length by computing the branching ratio *σ* for each subject, each neuronal avalanches and each time bin duration (Haldeman & Beggs, 2005). Indeed, a branching ratio ∼1 typically indicates a critical process. Specifically, the branching ratio was calculated as:

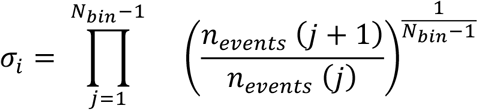

where σ is the branching parameter of the i-th avalanche in the dataset, Nbin is the total number of bins in the i-th avalanche, n_events_ (j) is the total number of events in the j-th bin. We then (geometrically) averaged the results over all avalanches as follow:

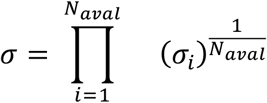

However, we repeated our analysis exploring different time bins, ranging from 1 to 5 obtaining similar results (see supplementary materials 2). For each avalanche, an *avalanche pattern* was defined as the set of all brain regions that were above threshold.

#### 2.5.2 Functional repertoire

The functional repertoire represents the number of unique avalanche patterns expressed during the recording (Sorrentino et al., 2021). *Unique* means that each avalanche pattern is counted only once to define the avalanche size of the functional repertoire (i.e., repetitions are discarded).

### 2.6 Transition matrices

We built an avalanche-specific transition matrix (ATM) in which its element (*i, j*) represented the probability that region *j* was active at time *t + δ*, given that region *i* was active at time *t*. The ATMs were averaged per participant and symmetrised (for more details see Sorrentino et al., 2021). We explored the frequency-specific ATMs. To this end, we filtered the source-reconstructed signal in the classical frequency bands (delta, 0.5–4 Hz; theta 4–8 Hz; alpha 8–13 Hz; beta 13–30 Hz; gamma 30–48 Hz). Thereafter, based on the frequency-specific ATMs, we calculated the Minimum Spanning Tree (MST) which was used, in turn, to estimate the topology of the avalanches and compare it between groups. In particular, we calculated both global and nodal topological parameters. The former were the *degree divergence* (*k*), which represents the amplitude of the degree distribution (i.e., how central the hubs are); the *leaf fraction* (*LF*) which is defined as the fraction of nodes with degree = 1; the *tree hierarchy* (*TH*), which conveys the trade-off between an efficiently connected network and its resiliency to targeted attacks, and the *assortativity*, that represent the tendency of a node to be connected to similar nodes in a given network (Jacini et al., 2018; Stam et al., 2014; Tewarie et al., 2015). Then, for what concerns the nodal parameters, we calculated the *Betweenness Centrality* (BC) and the *Degree*, which represents the topological importance of a node within the network (Rubinov & Sporns, 2010).

### 2.7 Multilinear regression analysis

We hypothesised that the alterations of brain dynamics could be used to predict the patients’ clinical impairment. Therefore, we built a multilinear regression model in which the neuropsychological tests represented the dependent variable while age, gender, education and brain dynamics features are the predictors. Multicollinearity was assessed through the variance inflation factor (VIF). To validate our approach, we performed *k*-fold cross-validation, with *k* = 5 (Varoquaux et al., 2017). Specifically, *k* iterations were performed to train our model and at each iteration the *k*^*th*^ subgroup was used as a test set.

### 2.8 Statistical analysis

Statistical analysis was carried out in MATLAB 2021a. The comparison between patients and HC was performed through permutation testing, shuffling the data labels (i.e., MCI or HC) 10.000 times. At each iteration, we recorded the absolute difference between the two randomly generated groups in order to generate a null distribution of absolute differences. Then, the actual absolute difference between patients and HC was compared to the random distribution to obtain a statistical significance. Finally, a possible relationship between the topological features of brain dynamics and the clinical conditions (hippocampal volumes and cognitive impairment) of the patients was explored through Spearman’s correlation. The results were corrected for multiple comparisons across both frequency bands and parameters using the False Discovery Rate (FDR) method (Benjamini and Hochberg, 1995) and the significance level was set at *p value* < 0.05 (corrected).

## 3. RESULTS

### 3.1 Cohort characteristics

As shown in table 1, the comparison between the demographic characteristics of the MCI and the HC groups revealed no significant differences in both age and education level. With respect to the structural information, only 30 MCI patients out of 32 and 22 out 32 HC completed the MRI scan. MCI patients exhibited a reduction of hippocampal volumes compared to the HC (p < 0.001). As expected, MCI displayed a worse cognitive performance. Specifically, the comparison between the neuropsychological evaluation of the two groups showed a significant difference in the: Mini Mental State Examination (MMSE) (p = 0.012), Rey’s 15-word immediate recall (p < 0.01), Rey’s 15-word delayed recall (p < 0.01), Word fluency (p < 0.01), Raven’s 47 progressive matrices (p < 0.01), Immediate visual memory (p < 0.01) and the FCSRT (immediate free recall, immediate total recall, delayed free recall, delayed total recall and index of sensitivity of cueing) (p < 0.001). No significant differences were found in the evaluation of the Beck Depression Inventory (BDI), Frontal Assessment Battery (FAB), Phrase construction, Freehand copying of drawings and Constructive apraxia with landmarks (see table 2).

### 3.2 Functional repertoire

In order to analyse the size of functional repertoire (i.e., the number of unique and different avalanches configurations in each participant), we used source reconstructed MEG data. As one can see in figure 2, the MCI group showed a reduction in the number of unique patterns in the theta band (*pFDR* = 0.047), revealing a restricted functional repertoire as compared to the HC.

**Figure 1:**
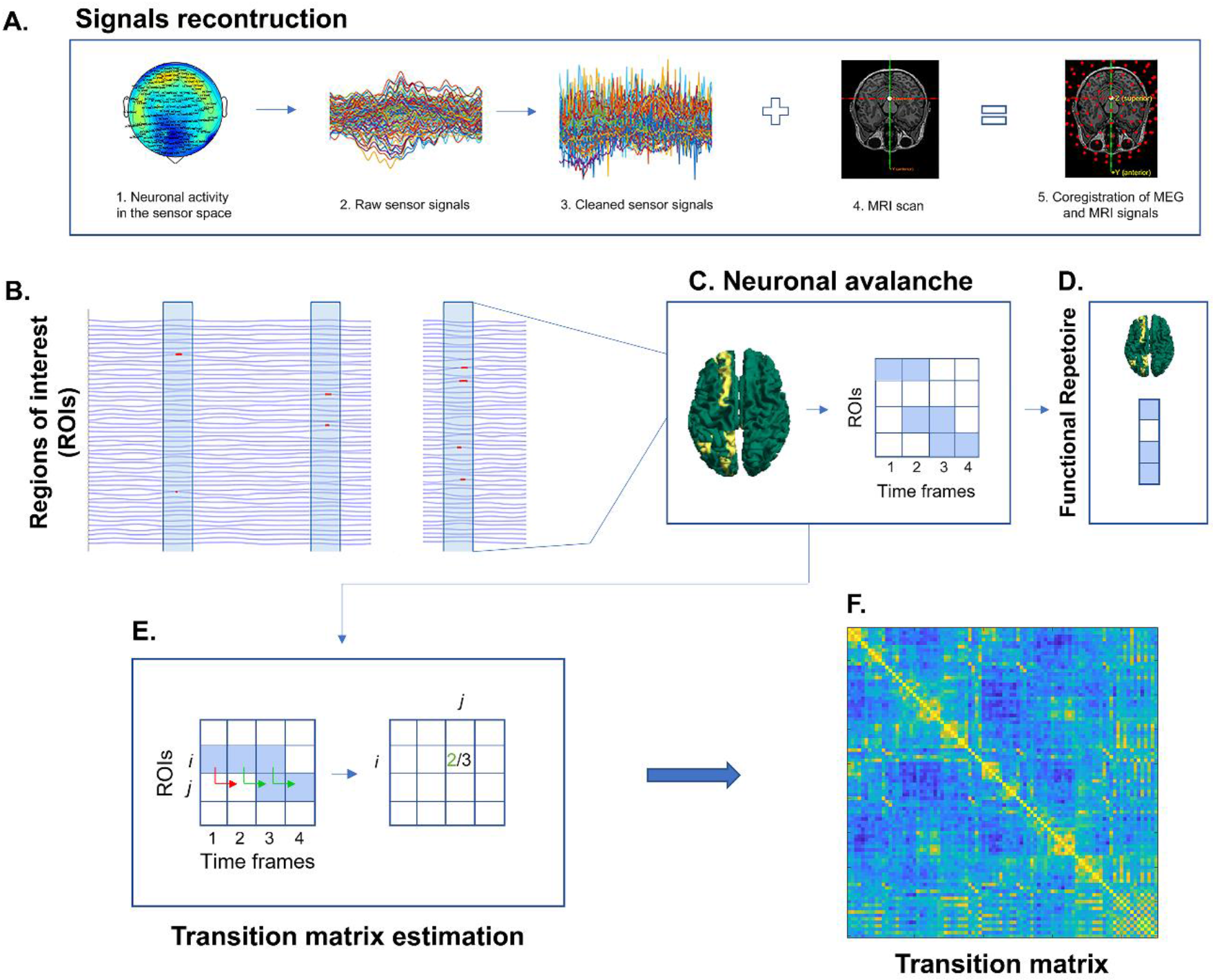
Pipeline overview. **(A)** 1. Registration of neuronal activity through magnetoencephalography (MEG); 2. Raw sensor signals including physiological artefacts; 3. Cleaned sensor signals (without physiological artefacts); 4. Magnetic resonance image (MRI) of the subject; 5. Beamforming obtained from the co-registration of MRI and MEG signals. **(B)** Source-reconstructed temporal series. The blue lines represent the z-score activity of a region, the light blue embossed boxes represent the time frame in which a neuronal avalanche occurred. Specifically, the red dots indicate the frame in which the time series was above threshold (z-score > 3). **(C)** Schematic representation of a neuronal avalanche. A neuronal avalanche occurred when at least one region was above threshold and ended when all the brain regions returned below threshold. In the brain plot, the areas above threshold are indicated in yellow while the ones below threshold are represented in green. On the right of the panel the avalanche is shown as a matrix. The ROIs are reported on the columns while the time frames are displayed on the rows. Blue boxes indicate the ROIs above threshold in each time frame. **(D)** Avalanche pattern representation. The blue boxes represent brain regions that were active during the avalanche. The totality of avalanche patterns occurring during a MEG scan represents the functional repertoire. **(E)** Schematic representation of a transition matrix. The light blue squares indicate that the region *i* was above threshold three times during the avalanche. Region *j* was active, after the activation of region *i*, only in two cases out of the three taken into consideration (as indicated by the green arrows). Thus, the probability that region *j* was active after the activation of region *i* was 2/3. **(F)** Average transition matrix.

**Figure 2:**
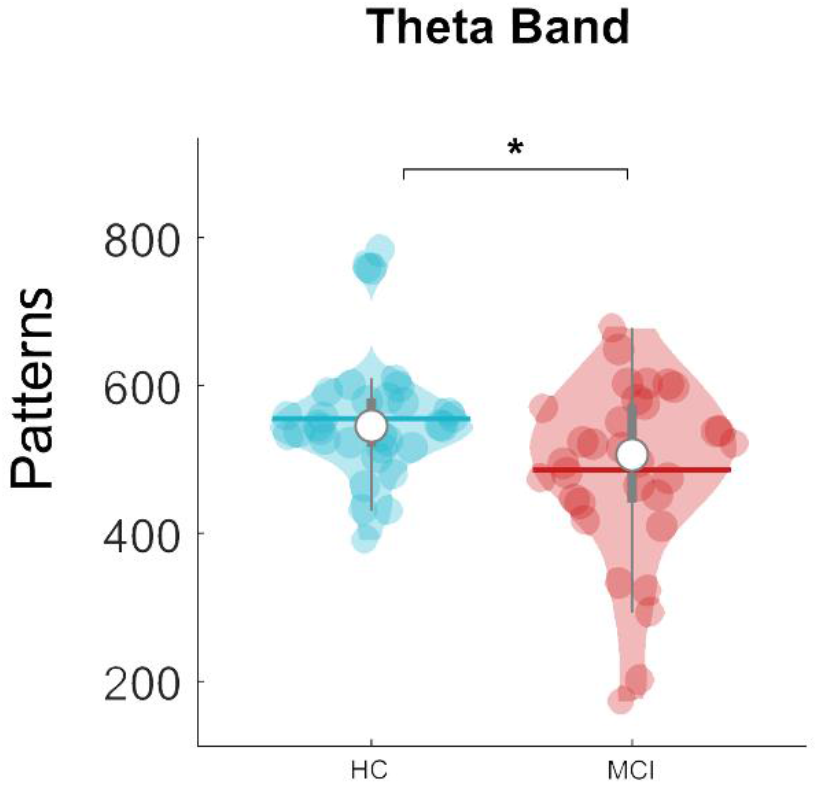
Functional repertoire comparison. The violin plots represent the comparison of the number of unique avalanche patterns between HC and MCI patients in theta band. MCI patients display a reduced functional repertoire compared to the HC. The median is represented by the white dot, the grey bar in the middle of the violins shows the first and the third quartile while the thin grey line is representative of the 95% confidence interval. The outliers are individually represented by the single dots. HC = healthy controls; MCI = Mild Cognitive Impairment; * p < 0.05.

### 3.3 Transition matrices topology

To study avalanche topology, we built an avalanche-specific transition matrix (ATM) (P. Sorrentino et al., 2021) for each subject. Then, we computed the minimum spanning tree for each ATMs (Wijk et al., 2010). Patients showed higher values of *LF* and *k* compared to the healthy subjects (Figure 3). Specifically, with respect to the *LF*, we found a statistically significant difference between the two groups in the delta (*pFDR* = 0.020) and theta bands (*pFDR* = 0.041). Similarly, we found a statistically significant difference of the *k* between the two groups in the delta (*pFDR* = 0.006), theta (*pFDR* = 0.046) and gamma band (*pFDR* = 0.020). We did not find any statistical difference neither for the nodal parameters (i.e., betweenness centrality and degree) nor for the remaining global parameter (i.e., the tree hierarchy).

**Figure 3:**
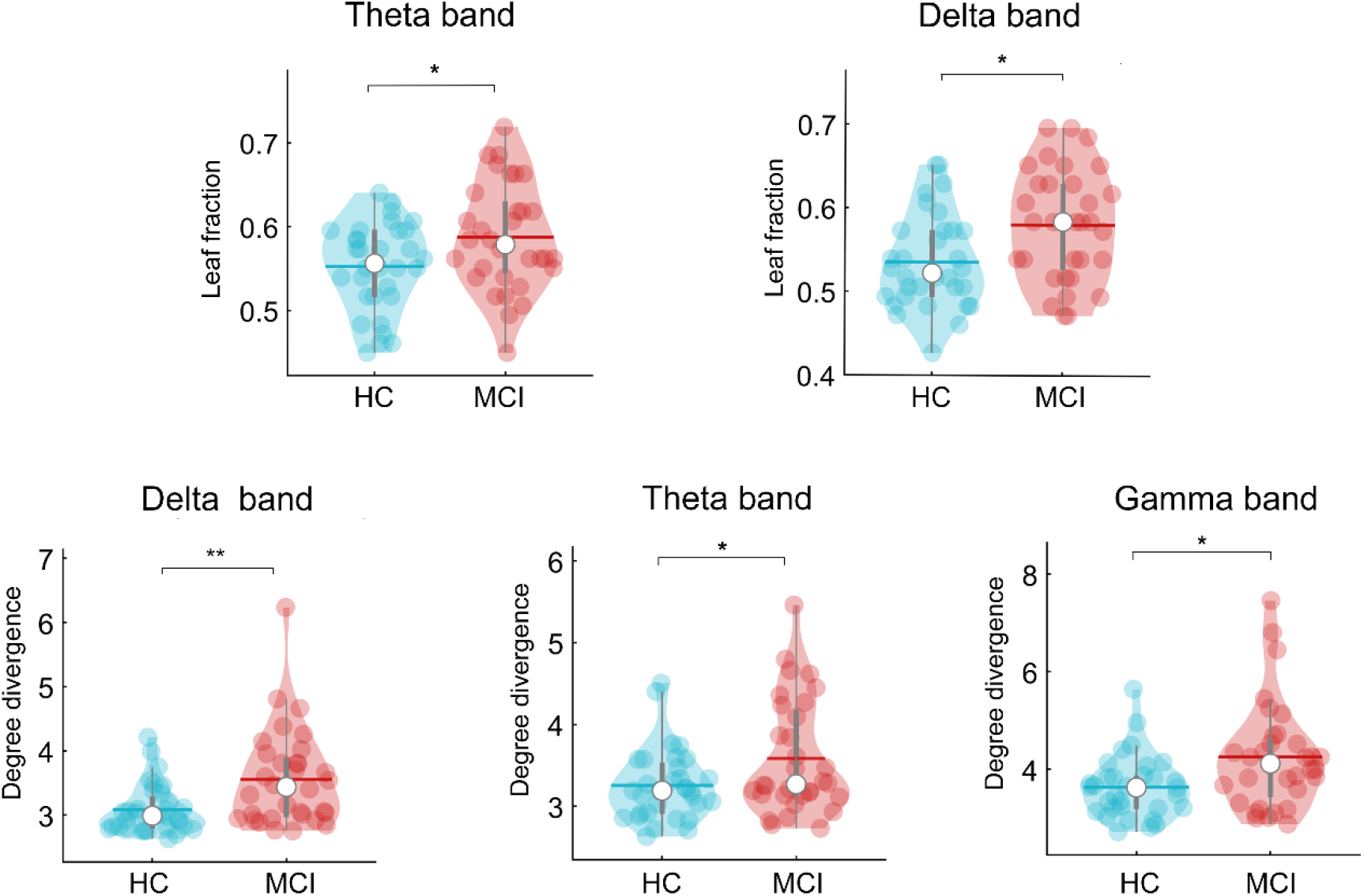
Topological parameters comparison. Violin plots represent the comparison between HC and MCI patients for topological parameters obtained from the transition matrices. MCI patients display higher values of leaf fraction in delta and theta band with respect to HC as well as higher values of degree divergence *(k)* in delta, theta and gamma band. *p < 0.05; **p < 0.01.

### 3.4 Correlations with clinical parameters

Spearman’s correlation was used to determine whether there was a relationship between topological network features and both the episodic memory (assessed through the Free and Cued Selective Reminding Test) and the structural parameters (i.e., hippocampal volumes). We focused on the FCSRT due to its sensitivity in detecting the hippocampal damage and amnesic deficits even in the earliest phase of disease (Auriacombe et al., 2010; Dubois et al., 2014). Firstly, we found a significant positive correlation, after FDR correction, between the *k* in the theta band and the FCSRT immediate total recall (*r* = 0.536; *pFDR* = 0.046). In addition, the *k* in the theta band positively correlated with the right hippocampal volume (*r* = 0.438; *pFDR =* 0.030) (Figure 4). Lastly, we found a significant positive correlation between the FCSRT immediate total recall and both right (*r* = 0.553; *pFDR* = 0.001) and left (*r=* 0.463; *pFDR=* 0.009) hippocampal volumes (data not shown). No significant correlation was found between the topological parameters and the other neuropsychological test scores and the left hippocampal volume.

**Figure 4:**
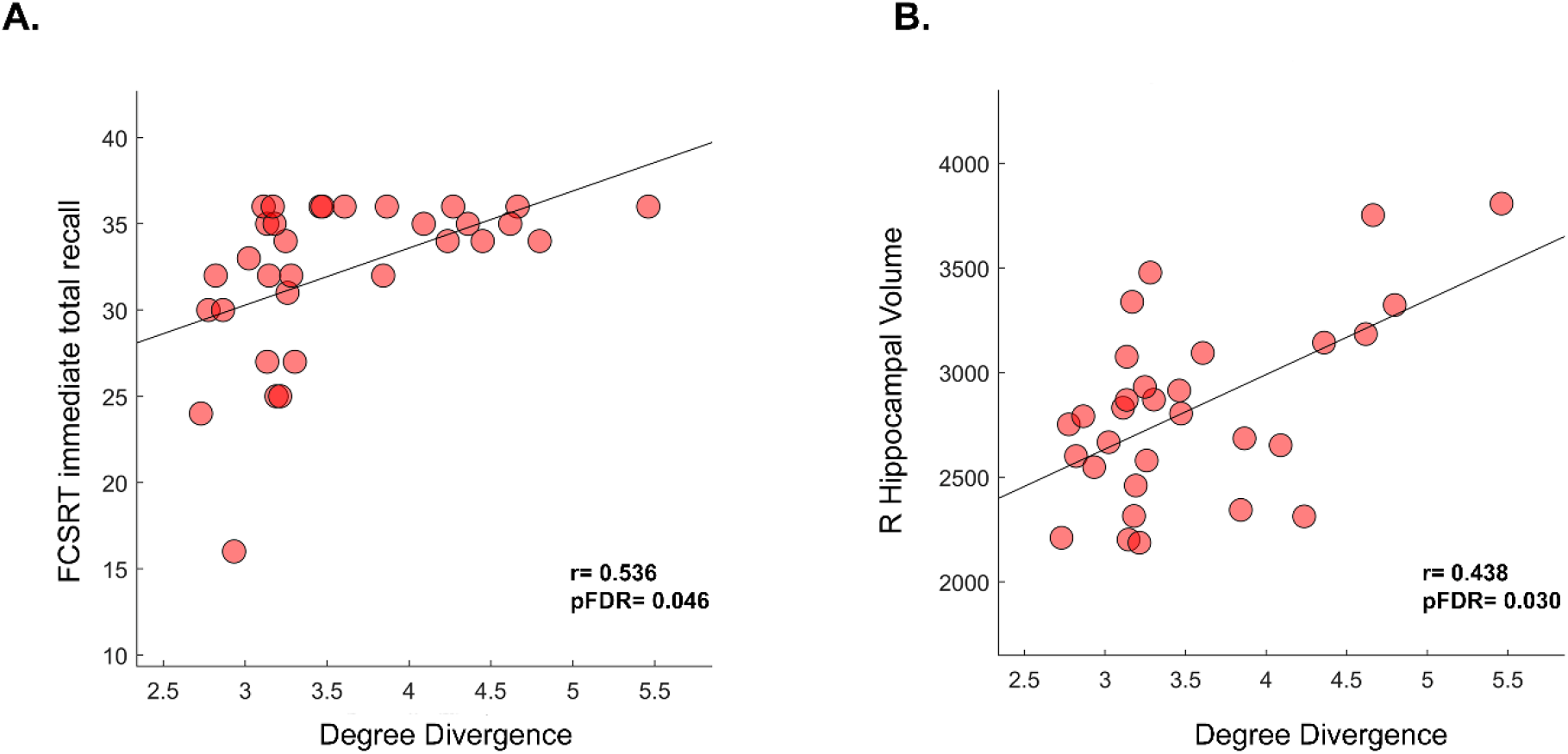
Correlation between MCI clinical features and degree divergence. (A) Spearman’s correlation between the *k* and the FCSRT immediate total recall. The positive correlation coefficient shows that as the *k* increases, the FCSRT score increases as well. (B) Positive correlation between the *k* and the right hippocampal volume. Higher values of *k* correspond to a greater preservation of the right hippocampal volume of the patients. For panel B the Spearman’s correlation analysis was performed taking into account the only 30 MCI patients who complete the MRI scan.

### 3.5 Predictive model: multilinear regression analysis

Taken into consideration the relationship between the *k* (i.e., the width of the degree distribution) and both the memory impairment and the right hippocampal atrophy, we wondered whether the width of the degree distribution could improve the prediction of the clinical impairment (as measured via the FCSRT immediate total recall). To this end, we built a multilinear regression model and we validated it using a k-fold cross validation approach. The model also contained four nuisance predictors i.e., age, education level, gender and the right hippocampal volume (see figure 5). We added the right hippocampal volume as a predictor given its role in memory and its involvement in MCI and AD (Apostolova et al., 2006; Devanand et al., 2012; Sarazin et al., 2010; Yavuz et al., 2007; Zammit et al., 2017). Our aim was to observe whether the *k* improved the prediction over the FCSRT immediate total recall as compared to using demographics and structural changes (i.e., right hippocampal volume) alone. We found that the *k* in the theta band significantly predicted the FCSRT immediate total recall (p = 0.041; R^2^ = 0.47; β = 2.7). Furthermore, the only other significant predictor was the education level (p = 0.012; β = 0.448) (figure 5). As an extra check, we re-built the same predictive model, but this time we switched the order of the last two predictors (i.e., the *k* and the right hippocampal volume). Our aim was to visually assess the residual predictive power of the hippocampus after the degree divergence was added. As shown in the supplementary materials (figure 4), the hippocampal volume did not significantly contribute to the prediction of the functional impairment after the functional degree divergence was taken into account.

**Figure 5:**
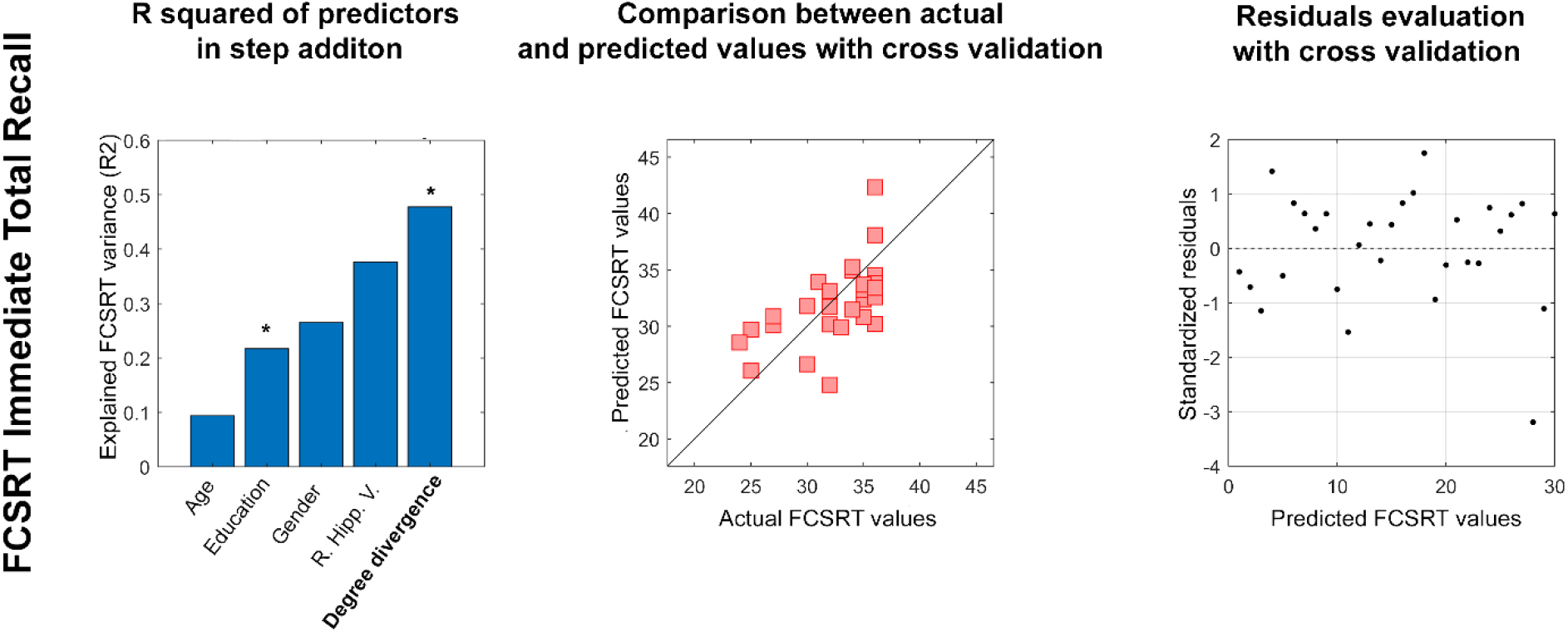
Clinical impairment prediction in the theta band. Multilinear regression analysis with *k-fold* cross validation was performed to verify the ability of the degree divergence (*k)* to predict the memory impairment assessed by the FCSRT immediate total recall. The left column displays the explained variance obtained by adding the predictors (age, education level, gender, right hippocampal volume and the *k* in the theta band). The significant predictors are highlighted in bold while the significant *p*-value is indicated with * (p < 0.05), **(p< 0.01). The central column displays the comparison between the predicted and the actual values of the responsive variable validated through the *k-*fold cross validation. Lastly, the right column shows the distribution of residuals which represent the standardisation of the difference between the actual and predicted values for the FCSRT immediate total recall.

## 4. DISCUSSION

In the present work we used source-reconstructed MEG data in a cohort of thirty-two MCI patients and thirty-two healthy subjects to explore the topological organisation of the large-scale brain dynamics, and its relationship with the clinical impairment. Firstly, we hypothesised that MCI subjects would show more stereotyped brain dynamics and, consequently, such stereotyped brain dynamics might result in a reorganisation of the spreading of the large-scale perturbations, as captured by changes in the functional topology. We used neuronal avalanches to quantify the flexibility of fast brain dynamics (Sorrentino, et al., 2021). The brain flexibility is given by its ability to generate a large number of functional configurations, that likely conveys a large number of different ways by which brain regions can interact among themselves (Shine et al., 2016; Zalesky et al., 2014). These large scale interactions manifest themselves as bursty activity (i.e., neuronal avalanches), and the number of different ways neuronal avalanches propagate across the brain defines the size of the functional repertoire (Chialvo, 2010; Sorrentino et al., 2021).

Our results showed that MCI patients exhibited a restricted functional repertoire (i.e., lower number of unique avalanche patterns) in the theta band compared to HC. Thus, MCI patients displayed a reduction of their brain flexibility which implies less reconfigurations of brain activity and a more stereotyped brain dynamics (Sorrentino et al., 2021). This is in line with previous reports that used the same approach in other neurodegenerative diseases such as Parkinson’s disease (Sorrentino et al., 2021) and Amyotrophic Lateral Sclerosis (Polverino et al., 2022), suggesting that the loss of flexibility might be a common dynamical change induced by different neurodegenerative processes.

Then, we generated the ATMs which provide information about the spatio-temporal avalanches dynamics (on the milli-second scale) estimating the probability that two brain regions will move away from their baseline activity in two consecutive time epochs (Sorrentino et al., 2021). By applying the MST on the ATMs, we proceeded to investigate their topological properties. The comparison between MCI patients and HC revealed that the former exhibited higher values of both *LF* (in the delta and theta band) and *k* (in the theta and gamma band) with respect to the latter. Higher *LF* values (i.e., fraction of nodes with degree = 1) are indicative of a more integrated network (as in a star-like topology), suggesting the idea that when the *LF* increases in the patients, the network shifts towards a more centralised organisation (Boersma et al., 2013). The *k* (i.e., the width of degree distribution) is a measure of the network’s synchronisability and its resilience against pathological events (Tewarie et al., 2015). Higher values of *k* convey presence of high degree nodes (Boersma et al., 2013) which, again, is consistent with a more centralised topology of the network. In line with previous evidence (Sorrentino et al., 2018), one might speculate that the higher centrality might be due to hubs compensating for the impairment of more peripheral nodes. As a consequence, the communication between brain regions becomes less efficient. Moreover, as the activations are forced to propagate through the hubs, this might indeed result in a less varied dynamics. However, these remarks remain purely speculative.

What stated so far is in line with our results showing that an increase of the *k* values reflects a better cognitive performance and is also correlated with greater right hippocampal volume. Indeed, the higher the *k* the better the FCSRT immediate total recall scores and the larger the hippocampal volume. A possible explanation might be that the less-compromised MCI subjects can put in place more effective compensation mechanisms, whereby the reduction of lower degree nodes might be effectively compensated by the more central hubs (Rucco et al., 2019). This view is consistent with the better clinical condition captured by both the higher FCSRT immediate total recall scores and the larger right hippocampal volumes. In addition, we found a positive correlation between the FCSRT immediate total recall scores and the right and left hippocampal volumes. Our results are in line with previous evidence (Jacini et al., 2018; Wang et al., 2006) which demonstrated the relationship between the loss of memory efficiency and hippocampal atrophy. For instance, a longitudinal study conducted by Stoub et al., showed that, over five years, MCI patients showed a progressive cognitive impairment directly related to changes of hippocampal volume (Stoub et al., 2010). Specifically, our results revealed a correlation between the topology (i.e., degree divergence in theta band) of the MCI and the right hippocampal volume. According to Cai and colleagues MCI patients show a reduction of their functional connectivity in the right hippocampus (which is also characterised by a greater rate of atrophy with respect to the left one). Importantly these alterations may be related to a worse cognitive performance expressed by a deficit of the episodic memory retrieval (Cai et al., 2017). This is in agreement with Heckers et al., who showed that the best retrieval of encoded words through a semantic cue was associated with higher right hippocampal activation (Heckers et al., 2002). However, we would like to specify that the brain network alterations in MCI are not only constrained to the right hippocampus compromission, but are often extended to the left hemisphere, reflecting, as well as the right hemisphere, a poorer cognitive performance. (Bai et al., 2009)

Noteworthy, in the current paper we wish to take advantage of the information about the fast brain dynamics, in order to further improve the individual predictions of clinical impairment (Bosboom et al., 2006; Chen et al., 2021). To this end, we built a multilinear regression model which demonstrated the predictive power of the degree divergence in the theta band over the FCSRT immediate total recall. Including the *k* in the model improved predictions over other nuisance predictors such as age, gender, education level and the right hippocampal volume. Interestingly, as shown in supplementary materials (figure 4) the structural information alone is unable to predict the clinical impairment (as assessed by the FCSRT immediate total recall), while functional rearrangement (defined by the higher degree divergence) carries predictive power. One possible explanation might be that the functional information provides a more comprehensive information, which also takes into account potential compensatory mechanisms which might be overlooked when focusing purely on structural data (i.e., atrophy). These findings highlight the importance of dynamical features to predict functional abilities (i.e., behaviour). Besides the *k*, only the education level resulted as a significant predictor for the FCSRT immediate total recall. Although the FCSRT scores are corrected by taking into account the education level, it is quite obvious that such a relationship would exist since it is expected that highly educated subjects are also those who exhibit better cognitive performance (Frasson et al., 2011). Finally, it is worth mentioning that the topological properties of both MCI patients and healthy subjects were obtained starting from the ATMs, that characterise the spatio-temporal evolution of neuronal avalanches. In other words, our evidence corroborates the clinical and behavioural relevance of scale-free, aperiodic activity at the fast time-scales.

## 5. CONCLUSION

In the current work we applied the novel framework based on avalanche transition matrices (ATMs) to investigate the spatio-temporal dynamics of neuronal avalanches in MCI and HC. As we hypothesised, MCI patients displayed a reduction of their brain dynamic flexibility which resulted in suboptimal functional topology and predicted the memory impairment. Based on these findings, we hope that the proposed framework may be helpful in monitoring the development of the disease and to effectively add subtle information derived from dynamical analysis to contribute to a more thorough phenotypical assessment of patients.

## Data Availability

The MEG data and the reconstructed avalanches are available upon request to the corresponding author, conditional on appropriate ethics approval at the local site. The availability of the data was not previously included in the ethical approval, and therefore data cannot be shared directly. In case data are requested, the corresponding author will request an amendment to the local ethical committee.

## Supplementary materials

**Figure 1:**
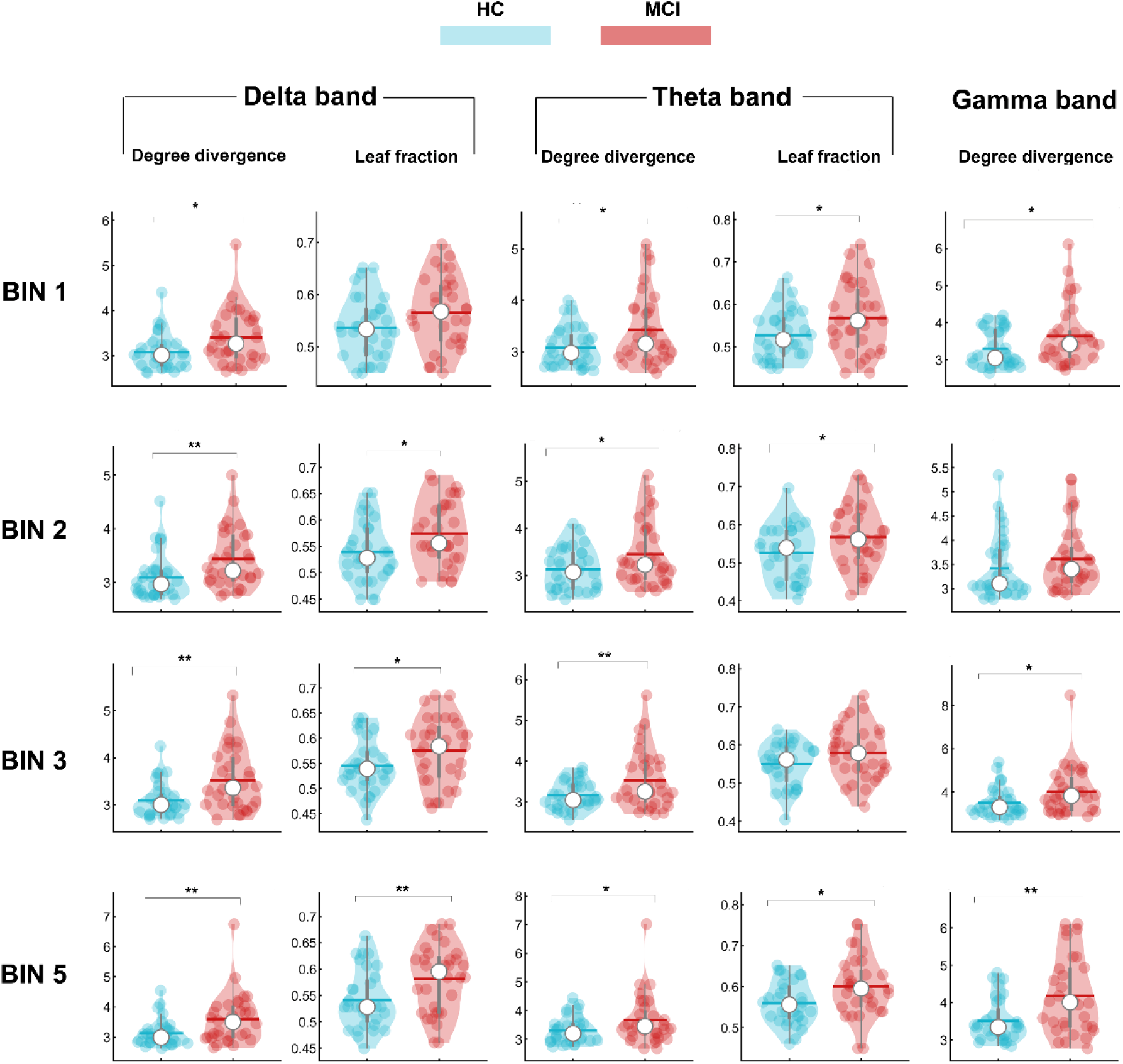
Topological parameters comparison across different binning. Violin plots represent the comparison between HC and MCI patients for topological parameters obtained from the transition matrices. For binning = 1, MCI patients display higher values of *k* in delta (p= 0.011), theta (p=0.012) and gamma band (p= 0.047) compared to HC, as well as higher values of Leaf fraction in theta band (p= 0.024). For binning =2, MCI patients display higher values of *k* and Leaf fraction in delta (p=0.007; p=0.025) and theta band (p=0.027; p=0.033) respectively. No statistical significance is observed for the *k* in gamma band. For binning=3, a statistical difference is observed for the *k* in delta (p=0.001), theta (p= 0.009) and gamma band (p=0.026). Concerning the leaf fraction MCI display higher values with respect to the HC in delta band (p= 0.048), while no statistical difference is observed for the Leaf fraction in theta band. Lastly, for binning=5 MCI patients display higher values of *k* in delta (p= 0.003), theta (p=0.030) and gamma band (p= 0.002) compared to HC, as well as higher values of Leaf fraction in delta band (p= 0.009) and theta band (p= 0.015).

**Figure 2:**
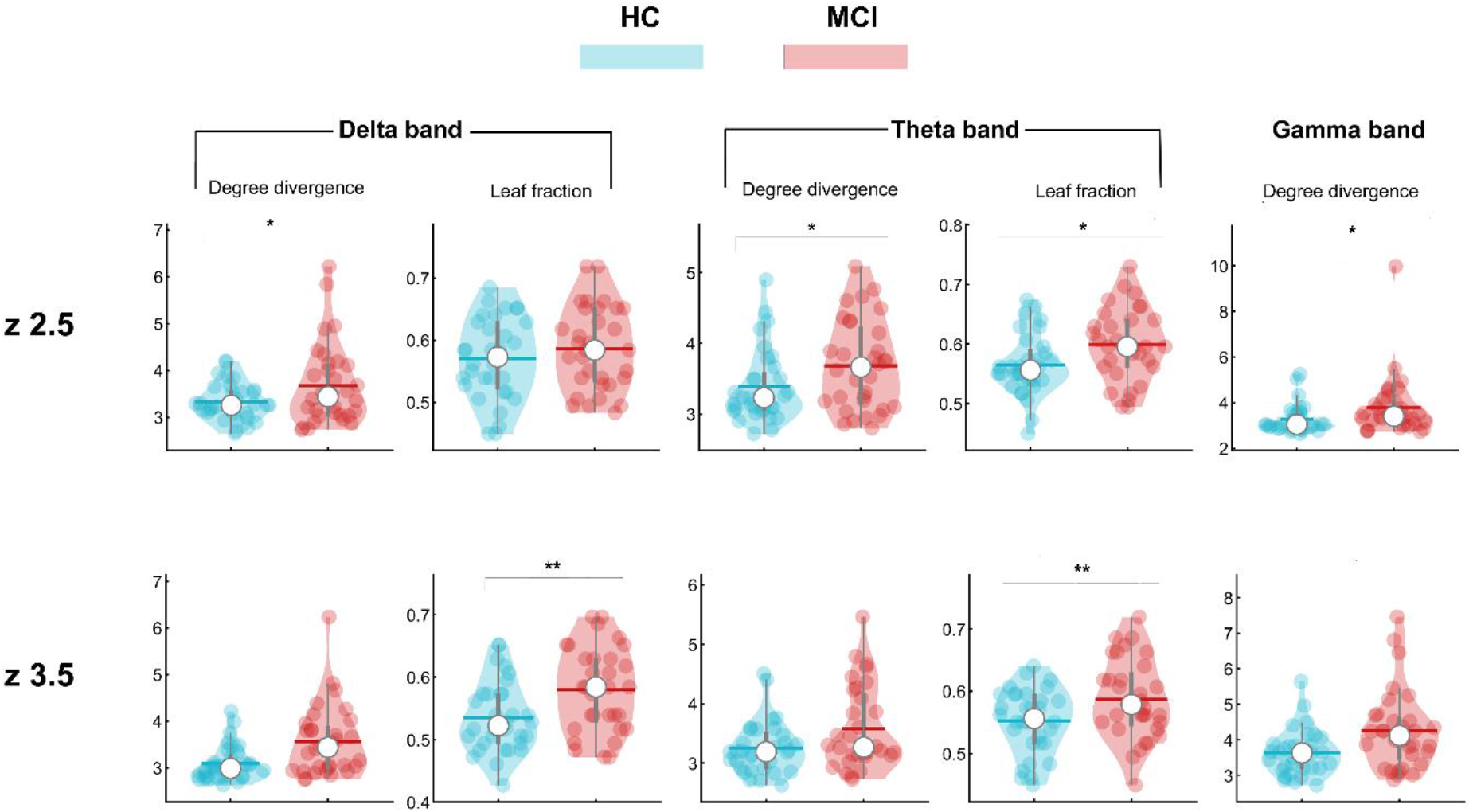
Topological parameters comparison across different thresholds. Violin plots represent the comparison between HC and MCI patients for topological parameters obtained from the transition matrices. For treshold *(z)* =2.5, MCI patients display higher values of *k* in delta (p= 0.039), theta (p=0.044) and gamma band (p= 0.031) compared to HC, as well as higher values of Leaf fraction in theta band (p= 0.018). For *z=* 3.5, MCI patients display higher values of both *k* and leaf fraction in theta band (p=0.003; p=0.007) while no statistically significant difference is observed for none of the remaining parameters.

**Figure 3:**
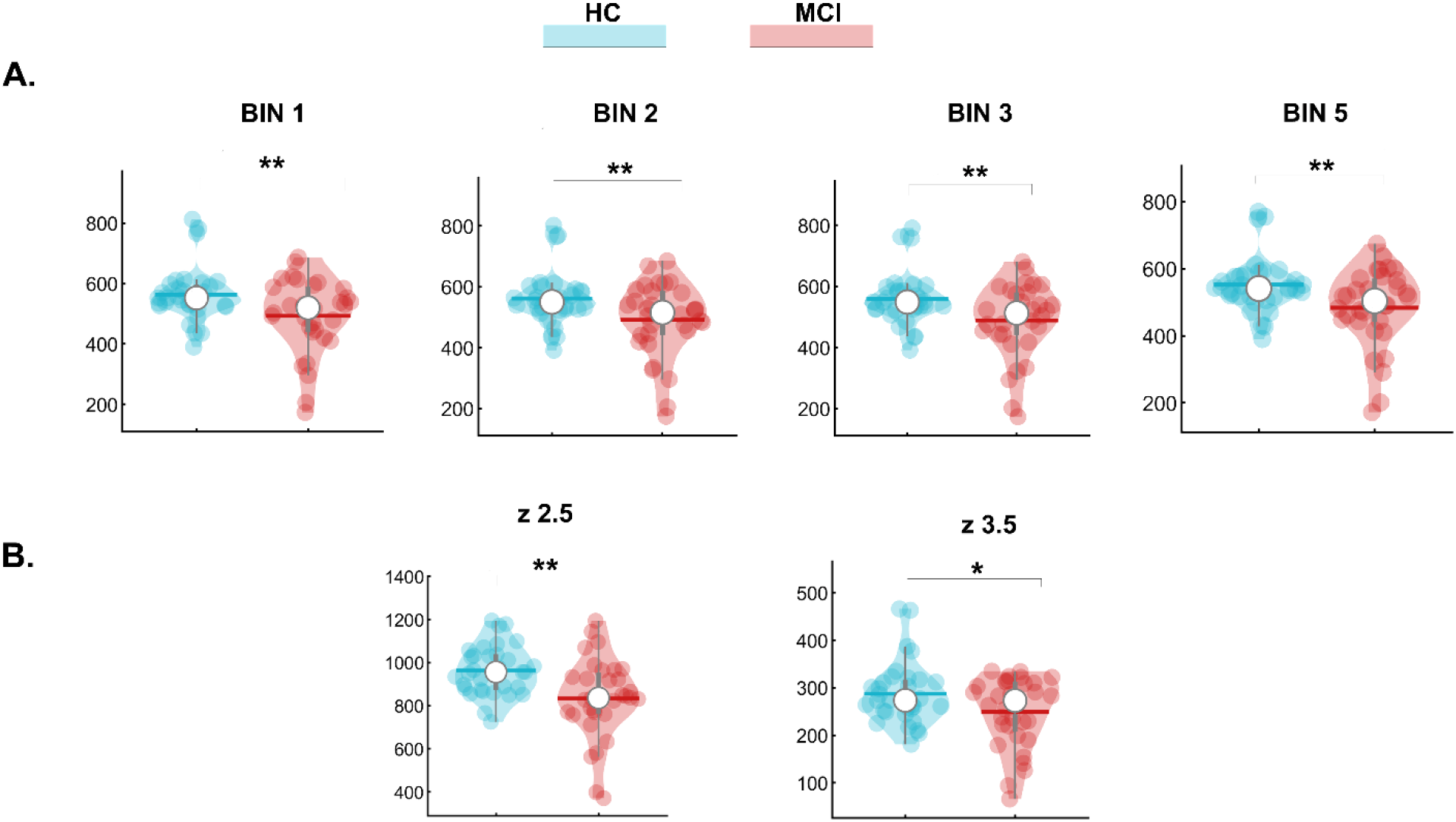
Functional repertoire comparison. (A) Violin plots representing the difference in the size of the functional repertoire in HC and MCI patients in theta band across different binning. For binning=1, p=0.009; for binning=2, p=0.010: for binning=3, p=0.006; for binning=5, p=0.008. (B) Violin plots representing the difference in the size of the functional repertoire in HC and MCI patients in theta band across different thresholds *(z)*. For *z=2*.*5*, p=0.001; for *z*=3.5, p=0.029.

**Figure 4:**
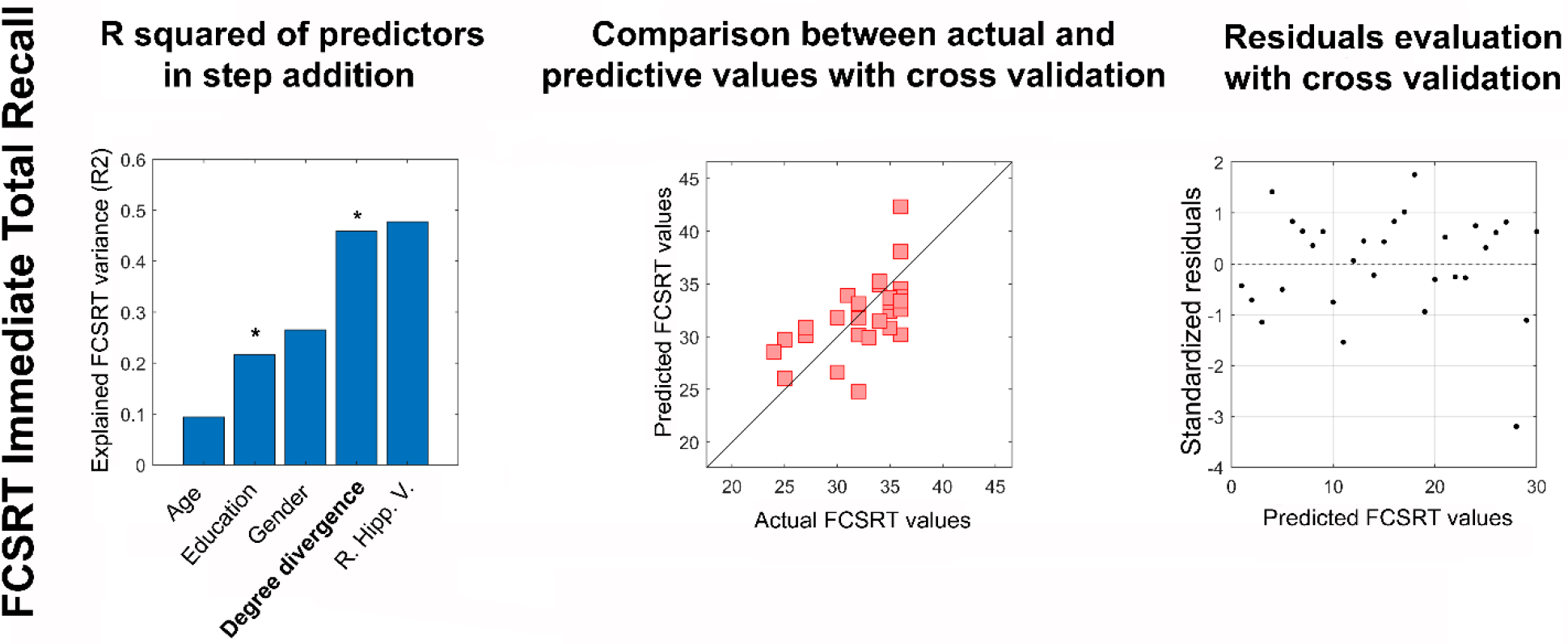
Clinical impairment prediction in theta band. Multilinear regression analysis with *k* fold cross validation. On the left column, we reported the explained variance obtained by adding the same predictor of figure 5 but switching the order of the two last predictors (i.e., the *k* and the right hippocampal volume) in order to visually assess how much variance was explained by the right hippocampal volume after adding all the other predictors. The significant predictor is highlighted in bold and the p value is indicated with * (p<0.05). The central column displays the comparison between the predicted and the actual values of the responsive variable validated through the *k* fold cross validation. Lastly, the right column shows the distribution of residuals which represent the standardisation of the difference between the actual and predicted values for the FCSRT immediate total recall.

## REFERENCES

Albert, M. S., DeKosky, S. T., Dickson, D., Dubois, B., Feldman, H. H., Fox, N. C., Gamst, A., Holtzman, D. M., Jagust, W. J., Petersen, R. C., Snyder, P. J., Carrillo, M. C., Thies, B., & Phelps, C. H. (2011). The diagnosis of mild cognitive impairment due to Alzheimer’s disease: Recommendations from the National Institute on Aging-Alzheimer’s Association workgroups on diagnostic guidelines for Alzheimer’s disease. Alzheimer’s & Dementia: The Journal of the Alzheimer’s Association, 7(3), 270–279. https://doi.org/10.1016/j.jalz.2011.03.008

Apostolova, L. G., Dutton, R. A., Dinov, I. D., Hayashi, K. M., Toga, A. W., Cummings, J. L., & Thompson, P. M. (2006). Conversion of Mild Cognitive Impairment to Alzheimer Disease Predicted by Hippocampal Atrophy Maps. Archives of Neurology, 63(5), 693–699. https://doi.org/10.1001/archneur.63.5.693

Auriacombe, S., Helmer, C., Amieva, H., Berr, C., Dubois, B., & Dartigues, J.-F. (2010). Validity of the Free and Cued Selective Reminding Test in predicting dementia: The 3C Study. Neurology, 74(22), 1760–1767. https://doi.org/10.1212/WNL.0b013e3181df0959

Bai, F., Zhang, Z., Watson, D. R., Yu, H., Shi, Y., Yuan, Y., Zang, Y., Zhu, C., & Qian, Y. (2009). Abnormal Functional Connectivity of Hippocampus During Episodic Memory Retrieval Processing Network in Amnestic Mild Cognitive Impairment. Biological Psychiatry, 65(11), 951–958. https://doi.org/10.1016/j.biopsych.2008.10.017

Boersma, M., Smit, D. J. A., Boomsma, D. I., De Geus, E. J. C., Delemarre-van de Waal, H. A., & Stam, C. J. (2013). Growing Trees in Child Brains: Graph Theoretical Analysis of Electroencephalography-Derived Minimum Spanning Tree in 5- and 7-Year-Old Children Reflects Brain Maturation. Brain Connectivity, 3(1), 50–60. https://doi.org/10.1089/brain.2012.0106

Bosboom, J. L. W., Stoffers, D., Stam, C. J., van Dijk, B. W., Verbunt, J., Berendse, H. W., & Wolters, E. Ch. (2006). Resting state oscillatory brain dynamics in Parkinson’s disease: An MEG study. Clinical Neurophysiology, 117(11), 2521–2531. https://doi.org/10.1016/j.clinph.2006.06.720

Buldú, J. M., Bajo, R., Maestú, F., Castellanos, N., Leyva, I., Gil, P., Sendiña-Nadal, I., Almendral, J. A., Nevado, A., del-Pozo, F., & Boccaletti, S. (2011). Reorganization of Functional Networks in Mild Cognitive Impairment. PLOS ONE, 6(5), e19584. https://doi.org/10.1371/journal.pone.0019584

Cai, S., Chong, T., Peng, Y., Shen, W., Li, J., von Deneen, K. M., Huang, L., & Alzheimer’s Disease Neuroimaging Initiative. (2017). Altered functional brain networks in amnestic mild cognitive impairment: A resting-state fMRI study. Brain Imaging and Behavior, 11(3), 619–631. https://doi.org/10.1007/s11682-016-9539-0

Carlesimo, G. A., Caltagirone, C., Gainotti, G., Fadda, L., Gallassi, R., Lorusso, S., Marfia, G., Marra, C., Nocentini, U., & Parnetti, L. (1996). The Mental Deterioration Battery: Normative Data, Diagnostic Reliability and Qualitative Analyses of Cognitive Impairment. European Neurology, 36(6), 378–384. https://doi.org/10.1159/000117297

Celone, K. A., Calhoun, V. D., Dickerson, B. C., Atri, A., Chua, E. F., Miller, S. L., DePeau, K., Rentz, D. M., Selkoe, D. J., Blacker, D., Albert, M. S., & Sperling, R. A. (2006). Alterations in Memory Networks in Mild Cognitive Impairment and Alzheimer’s Disease: An Independent Component Analysis. Journal of Neuroscience, 26(40), 10222–10231. https://doi.org/10.1523/JNEUROSCI.2250-06.2006

Chen, H.-J., Zou, Z.-Y., Zhang, X.-H., Shi, J.-Y., Huang, N.-X., & Lin, Y.-J. (2021). Dynamic Changes in Functional Network Connectivity Involving Amyotrophic Lateral Sclerosis and Its Correlation With Disease Severity. Journal of Magnetic Resonance Imaging, 54(1), 239–248. https://doi.org/10.1002/jmri.27521

Chialvo, D. R. (2010). Emergent complex neural dynamics. Nature Physics, 6(10), Article 10. https://doi.org/10.1038/nphys1803

Devanand, D. P., Bansal, R., Liu, J., Hao, X., Pradhaban, G., & Peterson, B. S. (2012). MRI hippocampal and entorhinal cortex mapping in predicting conversion to Alzheimer’s disease. NeuroImage, 60(3), 1622–1629. https://doi.org/10.1016/j.neuroimage.2012.01.075

Dubois, B., Feldman, H. H., Jacova, C., Hampel, H., Molinuevo, J. L., Blennow, K., DeKosky, S. T., Gauthier, S., Selkoe, D., Bateman, R., Cappa, S., Crutch, S., Engelborghs, S., Frisoni, G. B., Fox, N. C., Galasko, D., Habert, M.-O., Jicha, G. A., Nordberg, A., … Cummings, J. L. (2014). Advancing research diagnostic criteria for Alzheimer’s disease: The IWG-2 criteria. The Lancet Neurology, 13(6), 614–629. https://doi.org/10.1016/S1474-4422(14)70090-0

Fazekas, F., Chawluk, J. B., Alavi, A., Hurtig, H. I., & Zimmerman, R. A. (1987). MR Signal Abnormalities at 1.5 T in Alzheimer’s Dementia and Normal Aging. American Journal of Neuroradiology, 8(3), 421–426.

Folstein, M. F., Folstein, S. E., & McHugh, P. R. (1975). ‘Mini-mental state’. A practical method for grading the cognitive state of patients for the clinician. Journal of Psychiatric Research, 12(3), 189–198. https://doi.org/10.1016/0022-3956(75)90026-6

Fornito, A., Zalesky, A., & Breakspear, M. (2015). The connectomics of brain disorders. Nature Reviews Neuroscience, 16(3), Article 3. https://doi.org/10.1038/nrn3901

Frasson, P., Ghiretti, R., Catricalà, E., Pomati, S., Marcone, A., Parisi, L., Rossini, P. M., Cappa, S. F., Mariani, C., Vanacore, N., & Clerici, F. (2011). Free and cued selective reminding test: An Italian normative study. Neurological Sciences, 32(6), 1057–1062. https://doi.org/10.1007/s10072-011-0607-3

Friston, K. J. (2011). Functional and Effective Connectivity: A Review. Brain Connectivity, 1(1), 13–36. https://doi.org/10.1089/brain.2011.0008

Gong, G., He, Y., Concha, L., Lebel, C., Gross, D. W., Evans, A. C., & Beaulieu, C. (2009). Mapping anatomical connectivity patterns of human cerebral cortex using in vivo diffusion tensor imaging tractography. Cerebral Cortex, 19(3), 524–536.

Grober, E., & Buschke, H. (1987). Genuine memory deficits in dementia. Developmental Neuropsychology, 3(1), 13–36. https://doi.org/10.1080/87565648709540361

Haldeman, C., & Beggs, J. M. (2005). Critical Branching Captures Activity in Living Neural Networks and Maximizes the Number of Metastable States. Physical Review Letters, 94(5), 058101. https://doi.org/10.1103/PhysRevLett.94.058101

Heckers, S., Weiss, A. P., Alpert, N. M., & Schacter, D. L. (2002). Hippocampal and Brain Stem Activation during Word Retrieval after Repeated and Semantic Encoding. Cerebral Cortex, 12(9), 900–907. https://doi.org/10.1093/cercor/12.9.900

Ilardi, C. R., Chieffi, S., Scuotto, C., Gamboz, N., Galeone, F., Sannino, M., Garofalo, E., La Marra, M., Ronga, B., & Iavarone, A. (2022). The Frontal Assessment Battery 20 years later: Normative data for a shortened version (FAB15). Neurological Sciences, 43(3), 1709–1719. https://doi.org/10.1007/s10072-021-05544-0

Jacini, F., Sorrentino, P., Lardone, A., Rucco, R., Baselice, F., Cavaliere, C., Aiello, M., Orsini, M., Iavarone, A., Manzo, V., Carotenuto, A., Granata, C., Hillebrand, A., & Sorrentino, G. (2018). Amnestic Mild Cognitive Impairment Is Associated With Frequency-Specific Brain Network Alterations in Temporal Poles. Frontiers in Aging Neuroscience, 10. https://www.frontiersin.org/article/10.3389/fnagi.2018.00400

Liparoti, M., Troisi Lopez, E., Sarno, L., Rucco, R., Minino, R., Pesoli, M., Perruolo, G., Formisano, P., Lucidi, F., Sorrentino, G., & Sorrentino, P. (2021). Functional brain network topology across the menstrual cycle is estradiol dependent and correlates with individual well-being. Journal of Neuroscience Research, 99(9), 2271–2286. https://doi.org/10.1002/jnr.24898

Liu, Z., Zhang, Y., Yan, H., Bai, L., Dai, R., Wei, W., Zhong, C., Xue, T., Wang, H., Feng, Y., You, Y., Zhang, X., & Tian, J. (2012). Altered topological patterns of brain networks in mild cognitive impairment and Alzheimer’s disease: A resting-state fMRI study. Psychiatry Research: Neuroimaging, 202(2), 118–125. https://doi.org/10.1016/j.pscychresns.2012.03.002

López, M. E., Engels, M. M. A., van Straaten, E. C. W., Bajo, R., Delgado, M. L., Scheltens, P., Hillebrand, A., Stam, C. J., & Maestú, F. (2017). MEG Beamformer-Based Reconstructions of Functional Networks in Mild Cognitive Impairment. Frontiers in Aging Neuroscience, 9. https://www.frontiersin.org/article/10.3389/fnagi.2017.00107

Minati, L., Chan, D., Mastropasqua, C., Serra, L., Spanò, B., Marra, C., Caltagirone, C., Cercignani, M., & Bozzali, M. (2014). Widespread Alterations in Functional Brain Network Architecture in Amnestic Mild Cognitive Impairment. Journal of Alzheimer’s Disease, 40(1), 213–220. https://doi.org/10.3233/JAD-131766

Muñoz, M. A. (2018). Colloquium: Criticality and dynamical scaling in living systems. Reviews of Modern Physics, 90(3), 031001. https://doi.org/10.1103/RevModPhys.90.031001

Nolte, G. (2003). The magnetic lead field theorem in the quasi-static approximation and its use for magnetoencephalography forward calculation in realistic volume conductors. Physics in Medicine & Biology, 48(22), 3637.

Petersen, R. C. (2016). Mild Cognitive Impairment. Continuum : Lifelong Learning in Neurology, 22(2 Dementia), 404–418. https://doi.org/10.1212/CON.0000000000000313

Polverino, A., Lopez, E. T., Minino, R., Liparoti, M., Romano, A., Trojsi, F., Lucidi, F., Gollo, L., Jirsa, V., Sorrentino, G., & Sorrentino, P. (2022). Flexibility of Fast Brain Dynamics and Disease Severity in Amyotrophic Lateral Sclerosis. Neurology. https://doi.org/10.1212/WNL.0000000000201200

Romano, A., Trosi Lopez, E., Liparoti, M., Polverino, A., Minino, R., Trojsi, F., Bonavita, S., Mandolesi, L., Granata, C., Amico, E., Sorrentino, G., & Sorrentino, P. (2022). The progressive loss of brain network fingerprints in Amyotrophic Lateral Sclerosis predicts clinical impairment. NeuroImage: Clinical, 35, 103095. https://doi.org/10.1016/j.nicl.2022.103095

Rubinov, M., & Sporns, O. (2010). Complex network measures of brain connectivity: Uses and interpretations. NeuroImage, 52(3), 1059–1069. https://doi.org/10.1016/j.neuroimage.2009.10.003

Rucco, R., Lardone, A., Liparoti, M., Lopez, E. T., De Micco, R., Tessitore, A., Granata, C., Mandolesi, L., Sorrentino, G., & Sorrentino, P. (2022). Brain Networks and Cognitive Impairment in Parkinson’s Disease. Brain Connectivity, 12(5), 465–475. https://doi.org/10.1089/brain.2020.0985

Rucco, R., Liparoti, M., Jacini, F., Baselice, F., Antenora, A., De Michele, G., Criscuolo, C., Vettoliere, A., Mandolesi, L., Sorrentino, G., & Sorrentino, P. (2019). Mutations in the SPAST gene causing hereditary spastic paraplegia are related to global topological alterations in brain functional networks. Neurological Sciences, 40(5), 979–984. https://doi.org/10.1007/s10072-019-3725-y

Sarazin, M., Chauviré, V., Gerardin, E., Colliot, O., Kinkingnéhun, S., de Souza, L. C., Hugonot-Diener, L., Garnero, L., Lehéricy, S., Chupin, M., & Dubois, B. (2010). The Amnestic Syndrome of Hippocampal type in Alzheimer’s Disease: An MRI Study. Journal of Alzheimer’s Disease, 22(1), 285–294. https://doi.org/10.3233/JAD-2010-091150

Shine, J. M., Bissett, P. G., Bell, P. T., Koyejo, O., Balsters, J. H., Gorgolewski, K. J., Moodie, C. A., & Poldrack, R. A. (2016). The Dynamics of Functional Brain Networks: Integrated Network States during Cognitive Task Performance. Neuron, 92(2), 544–554. https://doi.org/10.1016/j.neuron.2016.09.018

Shriki, O., Alstott, J., Carver, F., Holroyd, T., Henson, R. N. A., Smith, M. L., Coppola, R., Bullmore, E., & Plenz, D. (2013). Neuronal Avalanches in the Resting MEG of the Human Brain. The Journal of Neuroscience, 33(16), 7079–7090. https://doi.org/10.1523/JNEUROSCI.4286-12.2013

Sica, C., & Ghisi, M. (2007). The Italian versions of the Beck Anxiety Inventory and the Beck Depression Inventory-II: Psychometric properties and discriminant power. In Leading-edge psychological tests and testing research (pp. 27–50). Nova Science Publishers.

Sorrentino, P., Rucco, R., Baselice, F., De Micco, R., Tessitore, A., Hillebrand, A., Mandolesi, L., Breakspear, M., Gollo, L. L., & Sorrentino, G. (2021). Flexible brain dynamics underpins complex behaviours as observed in Parkinson’s disease. Scientific Reports, 11(1), 1–12.

Sorrentino, P., Rucco, R., Jacini, F., Trojsi, F., Lardone, A., Baselice, F., Femiano, C., Santangelo, G., Granata, C., & Vettoliere, A. (2018). Brain functional networks become more connected as amyotrophic lateral sclerosis progresses: A source level magnetoencephalographic study. NeuroImage: Clinical, 20, 564–571.

Sorrentino, P., Rucco, R., Lardone, A., Liparoti, M., Troisi Lopez, E., Cavaliere, C., Soricelli, A., Jirsa, V., Sorrentino, G., & Amico, E. (2021). Clinical connectome fingerprints of cognitive decline. NeuroImage, 238, 118253. https://doi.org/10.1016/j.neuroimage.2021.118253

Sorrentino, P., Seguin, C., Rucco, R., Liparoti, M., Troisi Lopez, E., Bonavita, S., Quarantelli, M., Sorrentino, G., Jirsa, V., & Zalesky, A. (2021). The structural connectome constrains fast brain dynamics. ELife, 10, e67400. https://doi.org/10.7554/eLife.67400

Stam, C. J., Tewarie, P., Van Dellen, E., Van Straaten, E. C. W., Hillebrand, A., & Van Mieghem, P. (2014). The trees and the forest: Characterization of complex brain networks with minimum spanning trees. International Journal of Psychophysiology, 92(3), 129–138.

Stoub, T. R., Rogalski, E. J., Leurgans, S., Bennett, D. A., & deToledo-Morrell, L. (2010). Rate of entorhinal and hippocampal atrophy in incipient and mild AD: Relation to memory function. Neurobiology of Aging, 31(7), 1089–1098. https://doi.org/10.1016/j.neurobiolaging.2008.08.003

Tagliazucchi, E., Balenzuela, P., Fraiman, D., & Chialvo, D. (2012). Criticality in Large-Scale Brain fMRI Dynamics Unveiled by a Novel Point Process Analysis. Frontiers in Physiology, 3. https://www.frontiersin.org/article/10.3389/fphys.2012.00015

Tewarie, P., Prasse, B., Meier, J., Mandke, K., Warrington, S., Stam, C. J., Brookes, M. J., Van Mieghem, P., Sotiropoulos, S. N., & Hillebrand, A. (2022). Predicting time-resolved electrophysiological brain networks from structural eigenmodes. Human Brain Mapping, 43(14), 4475–4491. https://doi.org/10.1002/hbm.25967

Tewarie, P., van Dellen, E., Hillebrand, A., & Stam, C. J. (2015). The minimum spanning tree: An unbiased method for brain network analysis. NeuroImage, 104, 177–188. https://doi.org/10.1016/j.neuroimage.2014.10.015

Van Veen, B. D., Van Drongelen, W., Yuchtman, M., & Suzuki, A. (1997). Localization of brain electrical activity via linearly constrained minimum variance spatial filtering. IEEE Transactions on Biomedical Engineering, 44(9), 867–880.

Varoquaux, G., Raamana, P. R., Engemann, D. A., Hoyos-Idrobo, A., Schwartz, Y., & Thirion, B. (2017). Assessing and tuning brain decoders: Cross-validation, caveats, and guidelines. NeuroImage, 145, 166–179.

Wijk, B. C. M. van, Stam, C. J., & Daffertshofer, A. (2010). Comparing Brain Networks of Different Size and Connectivity Density Using Graph Theory. PLOS ONE, 5(10), e13701. https://doi.org/10.1371/journal.pone.0013701

Yavuz, B. B., Ariogul, S., Cankurtaran, M., Oguz, K. K., Halil, M., Dagli, N., & Cankurtaran, E. S. (2007). Hippocampal atrophy correlates with the severity of cognitive decline. International Psychogeriatrics, 19(4), 767–777. https://doi.org/10.1017/S1041610206004303

Zalesky, A., Fornito, A., Cocchi, L., Gollo, L. L., & Breakspear, M. (2014). Time-resolved resting-state brain networks. Proceedings of the National Academy of Sciences, 111(28), 10341–10346. https://doi.org/10.1073/pnas.1400181111

Zammit, A. R., Ezzati, A., Zimmerman, M. E., Lipton, R. B., Lipton, M. L., & Katz, M. J. (2017). Roles of hippocampal subfields in verbal and visual episodic memory. Behavioural Brain Research, 317, 157–162. https://doi.org/10.1016/j.bbr.2016.09.038

